# Acceptability and gut-modulatory effects of Finger millet-based complementary food (FMCF) in treating Indian children with Moderate Acute Malnutrition: A randomized controlled trial protocol

**DOI:** 10.1101/2025.11.07.25339662

**Authors:** Sakshi Rai, Nikhita B. Ramesh, Parth Sarin, Sahil Bipin Kumar Suthar, Akshay Shinde, Santosh Kumar Banjara, Aruna V. Reddy, G Vijayalakshmi, Hemant Mahajan, Ananthan Rajendran, R Naveen Kumar, Paras Sharma, Sudhakar Ajmera, Deepak Sharma, S Radhika Madhari, Sourav Sengupta, Devaraj J. Parasannanavar

**Author notes:** Corresponding Authors: Devaraj J. Parasannanavar, Scientist D, Maternal and Child Health Nutrition Division, ICMR-National Institute of Nutrition, Hyderabad. Email ID, Contact: 8074928620. Sakshi Rai and Nikhita B. Ramesh have contributed equally and shared first authorship.

## Abstract

**Background:** Undernutrition among children under five remains a critical global health challenge, contributing to higher mortality. Traditional therapeutic foods often overlook the gut microbial derangements, critical for nutrient assimilation, growth, and long-term recovery. Finger millet, with its exceptional nutritional profile, stands as a sustainable alternative to cereal-based foods to address this nutritional gap while supporting gut-health and inflammation.

**Objectives:** The aim of this study is to evaluate a Finger Millet-based Complementary Food (FMCF) as a nutrient-dense, safe, and acceptable intervention for moderately acute malnourished (MAM) children.

**Methods:** Finger millet (GPU-28) was germinated, roasted, and gamma-irradiated to improve nutrient bioavailability, reduce antinutrients, and ensure microbial-safety. We enrolled 200 children (100 MAM, 100 Healthy controls) aged 18-59 months from Anganwadi Centres. Participants have been randomized to receive either FMCF or an equicaloric wheat-based control (Balamrutham+) for 8-weeks. Anthropometry, dietary intake, gut-microbiota composition, and inflammation marker estimation and correlation will be performed at different timepoints.

**Results:** Product analysis revealed superior protein-energy ratio, fibre, calcium, and micronutrient content in FMCF compared to Balamrutham+. Microbiological and aflatoxin testing confirmed safety and shelf-life stability. Pilot feeding trial showed higher consumption (81–98%) and shorter eating times (53–71%, p<0.001).

**Conclusion:** FMCF is promising, cost-effective, and scalable intervention for nutritional-recovery in MAM, with potential to modulate the gut-microbiome and alleviate inflammation. The product aligns with SDG-2 (Zero Hunger) and SDG-3 (Good Health and Well-being), reinforcing its relevance as a public-health intervention.

**Approval and registration:** Institutional Ethics Committee (IEC)-CR/2/V/2023 and Clinical Trials Registry of India (CTRI)-CTRI/2023/06/053590.

## 1. Introduction

Malnutrition is a significant global health burden that has been prevailing in Low- and middle-income countries (LMICs), accounting for one in every five deaths (1). The origins of this condition are multifactorial, often arising from inadequate dietary intake, poor nutrient absorption, frequent infections, and a disrupted gut microbiota that affects metabolic function (2). If left unattended, these nutritional deficiencies can irreversibly alter the developmental trajectories, impairing immunological, cognitive, and physical development (3). Malnutrition not only impacts immediate health outcomes but also increases the DALYs (Disability Adjusted Life Years) (4).

According to the Joint Child Malnutrition Estimates (JME) 2025, the situation for children under 5 continues to be concerning, with 150.2 million stunted and 42.8 million wasted globally. Notably, South Asia alone accounts for 56.4 million stunted and 24.4 million wasted children. Referring to the latest stats, Africa and South Asia continue to rank highest in population of stunted and wasted children under 5, respectively (5), (6). To tackle this burden, various therapeutic feeding and nutritional intervention programs have been initiated worldwide. These nutritional programs are primarily designed to manage acute malnutrition, given its reversible nature (3). These policies are aimed at delivering a calorie-dense nutritional supplement to fulfil the daily requirements as per the Recommended Dietary Allowance (RDA). Programs like Ready-to-use Therapeutic Food (RUTF), Energy-Dense Nutritional Food (EDNF), Fortified Blended Food (FBF), Micronutrient Powders (MNPs), and Take-Home Ration (THR), were introduced. These policies also provide mid-day meals, preschool education, primary healthcare, immunisation, and health check-ups to the children (7), (8). The conventional RUTF treatment involved a one-week course of antibiotic administration prior to the feeding. The use of routine antibiotics before therapeutic feeding resulted in a higher recovery rate and a reduced mortality rate (9), (10). However, it is important to note that this treatment does not address the compromised status of the gut, which is an inherent condition among these groups (11). This leads to an immediate improvement of anthropometric parameters but results in a reversal to an undernourished state post-intervention. To validate this, a study conducted by the Gordon lab of Washington University in St. Louis, USA, discovered that treatment with RUTF was insufficient to persistently restore a balanced gut microbiota. The findings suggested the need for interventions with higher therapeutic efficacy on both nutritional and gut-microbial outcomes (12).

Available evidence from the literature highlights a strong association between gut microbiota and undernutrition. Distinct microbial signatures have revealed a compositional disparity in the gut microbiota of undernourished and healthy children (Ghosh, Sen Gupta, et al., 2014; Iddrisu et al., 2021b). An undernourished gut exhibits a decreased bacterial richness, increased abundance of pathogenic genera, and a loss of beneficial genera (13) (14). These alterations are likely to impair or disrupt the overall functional capacity.

Both acute and chronic malnutrition have distinct aetiologies and patterns of onset. Severe Acute Malnutrition (SAM), is associated with a reduction in *Bacteroides* and an increase in *Proteobacteria* (15). These shifts contribute to gastrointestinal disturbances, such as diarrhea and malabsorption, ultimately resulting in diminished nutrient uptake and weight loss. In contrast, chronic malnutrition, is linked to a reduced abundance of *Prevotella 9, Clostridia*, and a specific enrichment of Campylobacter, Akkermansia, and *Escherichia coli*, indicating altered gut ecology over a prolonged period (16), (17). Moreover, the undernourished gut has been found to have high proteolytic activity and is accompanied by a low threshold of enteropathogens. This collectively triggers a subclinical constellation of intestinal pathologies that include inflammation, barrier dysfunction, predilection to pathogen invasion, altered transit, and malabsorption (18)(19), (20) (21), (22). Gut inflammation induces production of anti-microbial peptides in response to pathogen growth, but can lead in depletion of the commensals, having an ecological impact in the gut (23). Inflammation results in increased luminal oxygen levels, diminishing numbers of strict anaerobes and promoting the growth of aero-tolerant microbes, especially pathogens groups eventually leading to a state of dysbiosis (24) (25) (26).

These facts warrant rethinking of strategies to tackle the burden of undernutrition and provide an overall improved food environment for children, ensuring a healthy childhood and optimum physical and mental growth. In this study, we aimed to develop nutritional intervention strategies that provide sufficient calories and can simultaneously restructure the gut microbiota. With the recent focus on millets, their exceptional nutritional profile and effective gut-modulatory potentials, we aim to pick them as our approach (27), (28). Talking about the previous times, millets were deeply embedded in the traditional diets of the Indian population, providing high nutritional value with their unique blend of dietary fibres and polyphenols. (29). Millets such as Sorghum and Pearl millet made up 40% of all grains cultivated before the Green Revolution, providing a diverse array of benefits. (30). With the decline in its consumption, this dietary shift resulted in nutritional imbalances. Restoring the use of these traditional crops for their benefits can give us an affordable and sustainable outcome. Finger millets, due to their exceptional nutrient profile, can be a promising candidate in one’s diet. High amounts of polyphenol in finger-millet grains have been shown to impart indirect anti-mutagenic, anti-glycemic, and anti-oxidative properties.(31), (32). Their proportion of dietary fibre in finger millet is also relatively higher than that of the routinely used cereals like refined wheat and maize. (33), (34). The fermentability of these soluble and insoluble dietary fibres in the large intestine by resident gut microbes can potentially drive the growth and/or activity of health-promoting bacteria in the gastrointestinal tract. This microbial fermentation generates a diverse range of bioactive molecules, termed metabolites. (35). This includes vitamins and short-chain fatty acids (SCFAs), such as butyrate and propionate, which not only serve as direct energy sources for colonic epithelial cells but also play a crucial role in activating various signalling pathways. (36). Activation of these pathways influences appetite control, inflammation, gut motility, and energy expenditure by increasing fatty acid oxidation and adaptive thermogenesis. SCFAs also have anti-inflammatory effects and can protect against oxidative stress. Hence, we hypothesise that the use of finger millets and the action of its numerous nutrient and non-nutrient components on the undernourished gut microbiome and the host will successfully resolve the multifaceted problems of undernutrition in order to restore the lost resilient factor/s as in healthy children.

**Objectives:** (1) develop a nutrient-dense, culturally accepted millet-based food, (2) test its efficacy in an RCT targeting undernourished children, (3) evaluate its potential in modulating the gut-microbiota of MAM children, (4) quantify changes in gut-derived and systemic inflammatory biomarkers to investigate immunomodulatory effects and assess long-term sustainability.

## 2. Materials and Methods

### 2.1. Study Design and Setting

#### 2.1.1 Study setting

This study is a community-based proof-of-concept clinical trial. We aimed to investigate the effect of a finger-millet-based dietary intervention compared to a wheat-based formulation on gut microbiota and nutritional health. The target study groups included cases and controls aged 18-59 months. Cases were classified as having primary moderate acute malnutrition [MAM: characterized by median weight-for-height z scores (WHZ) ranging from -3 to -2 SD according to WHO growth standards], and they were currently free of any co-morbidities. Conversely, controls were classified as apparently healthy children with WHZ scores between -1 and +1.

Children were recruited from Anganwadi Centres (AWCs) in Telangana, including areas around Hyderabad. This involved visiting over 200 AWCs and screening a total of 2398 children under 5 years old. Participants were then randomly assigned to either the FMD or BMD intervention group. Randomization was employed at the AWC level, not at the individual level, to ensure each Anganwadi centre received only one type of intervention. Controls were age and gender matched with cases, serving as a reference group, and did not receive any intervention.

#### 2.1.2 Study design

The study was approved by the Institutional Ethics Committee (IEC Approval No. CR/2/V/2023) and registered under the Clinical Trials Registry-India (CTRI/2023/06/053590). The study employs a longitudinal design and is conducted over a period of 6 months (i.e., 24 weeks). This 24-week period comprises five time points of data collection: 0 days, 4 weeks, 8 weeks, 16 weeks, and 24 weeks. As depicted in the figure_1, the feeding or intervention period spans 8 weeks. Following the intervention, two follow-up assessments were conducted at regular intervals (8 weeks each). Fecal samples and anthropometric data were collected at all five time points. In contrast, blood samples were taken at three specific times: the baseline (0 days), at the end of the feeding period (8th week), and during the second follow-up (24th week). For healthy participants, both fecal and blood samples were only collected at baseline.

A total of 200 children were planned for enrolment, including 100 moderately acutely malnourished (MAM) and their age- and gender-matched healthy children. These 100 MAM children were randomized into two intervention arms FMD and Bal-D. Respective intervention products were provided (60–150 g dry weight based on body weight) in the form of laddu, porridge, idly, or dosas as a morning and evening snack to the enrolled participants. Individual servings were calculated according to the guidelines of the Supervised Supplementary Feeding Program (SSFP). The dietary intervention was administered at least 5 days per week under supervision for a total of 8 weeks.

#### 2.1.3 Sample size

Sample size estimation for this study was performed using two complementary methods. For outcomes related to anthropometric changes, calculations were based on results from a pilot study conducted at AWCs, resulting in an estimated sample size of 50 participants per arm. Additionally, a microbiome-based approach was applied using reference data from Gordon et al. (2021). The following formula was used 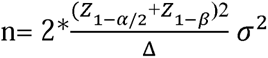, where Z*_1-a/2_* corresponds to the chosen significance level (e.g., 1.96 for α = 0.05), Z_β_ represents the desired statistical power (e.g., 0.84 for 80%), σ^2^ is the variance of the alpha diversity index from prior studies, and δ is the minimum detectable difference (effect size) between group means. An effect size of 0.68 and variance of 0.39, as reported by Gordon et al. (2021), were used, leading to a calculated sample size of 48 per group (total n = 96). The study design thus targeted this sample size to ensure adequate power for both anthropometric and microbiome outcomes.

### 2.2. Product Development Process

#### 2.2.1. Ingredient Selection and Procurement: Specify raw materials, origins (Finger millet GPU-28, dates)

The present investigation was conducted at the ICMR–National Institute of Nutrition, Department of Clinical Epidemiology, Hyderabad, Telangana, India. Finger millet (Eleusine coracana, GPU-28 variety) grains were procured from Karnataka and processed at a private facility in Bengaluru. The GPU-28 variety was selected for its widespread cultivation and availability in southern states such as Karnataka and Andhra Pradesh, making it a suitable choice for developing regionally relevant nutritional products. Dried date powder was sourced from Tamil Nadu, while other ingredients and formulations were supplied by Telangana Foods, a Government of Telangana enterprise based in Hyderabad.

#### 2.2.2. Processing Workflow: Describe soaking, germination, roasting, gamma irradiation with parameters (can attach flowchart in Supplementary)

##### Millet processing

The grains were cleaned, sorted, and washed with sterile ozonated water, and then soaked for 24 hours. After decanting the water, Germination was initiated by laying the soaked grains on a rectangular tray lined with a moist paper sheet and placed in low-light conditions. Water was sprayed every 4 hours to maintain optimal conditions for sprouting. The grains were allowed to germinate for 48 hours (37), (38), (39), (40), (41), (42). Afterwards, the sprouted grains were dried in a dehydrator at 50°C for 8 hours. The dried grains were then roasted in a drum roaster at 100–130°C for 60 minutes, until they became aromatic, crunchy, and developed their distinct earthen flavor. Roasting helped reduce the natural bitterness and enhanced the flavor (43). The roasted grains underwent milling and sieving, resulting in fine flour, which was stored in an airtight container in the refrigerator for later use in the preparation (Figure_2).

#### 2.2.3. Formulation Composition: Include a comparative table (FMCF vs. control RUTF/Balamrutham)

Finger millet–based Ready-to-Use Complementary Food (FMCF) was developed in line with the existing Balamrutham+ supplementary product, designed to provide ∼75 kcal per kg body weight per day for moderately acute malnourished children. This energy level is consistent with WHO/UNICEF recommendations of 70–100 kcal/kg/day for catch-up growth in MAM management and reflects the benchmark already established by ICMR-NIN formulations (44). The formulation was prepared in consultation with an expert group at ICMR–NIN, Hyderabad, Telangana, India.

##### Product F1-FMCF

To develop complementary food (FMCF), Germinated-roasted finger millet flour and date powder were used in the mix preparation. All the ingredients were roasted and proportioned as shown in Figure_3.

##### Product F2 - RUTF2

The existing product has been slightly modified by replacing sugar with a natural sweetener, Date powder. The ingredients and formulation are detailed in Figure_3.

###### Standardization of Recipes

The prepared mixture can transform into various recipes to add variety. Recipes prepared using these two mixes were standardized to yield a serving size of 30g each. Initially, various recipes including porridge, laddu, sweet-steamed cake (Idly), pan cakes, pan-fried dumplings, cake, and biscuits were tried and tested. Considering factors such as instant and easy preparation, shorter cooking time and alignment with the serving size, four of the above recipes were selected for intervention. Porridge, Laddu, Idly (sweet-steamed cake) and pancake were the ideal choices and suited both mixes perfectly. The preparation of these recipes is descripted in Figure_3 .

#### 2.2.4. Nutritional and Bioactive Profiling: AOAC methods, proximate, mineral, and antioxidant analyses

The proximate composition and dietary fibre content of the products were determined according to the standard procedures of the Association of Official Analytical Chemists (AOAC) (refer to Supplementary File). Mineral concentrations were quantified using Flame Atomic Absorption Spectroscopy (FAAS) following the AOAC (2005) protocol.

The bioactive components and antioxidant properties were analyzed through established spectrophotometric methods. Total Phenolic Content (TPC) was estimated using the Folin–Ciocalteu method described by Gao et al. (2002), while Total Flavonoid Content (TFC) was assessed following the method of Jia et al. (1998). The antioxidant activity was evaluated using the DPPH radical scavenging assay as modified from Brand-Williams et al. (1995). Detailed analytical procedures are presented in the Supplementary File.

#### 2.2.5. Safety and Shelf-Life Tests: Aflatoxin testing, microbial load, and gamma-irradiation validation results

Aflatoxin testing was carried out on finger millet flour and peanut samples, with analysis of aflatoxin B1 and B2 following the AOAC method (45). The method had a limit of detection (LOD) of 1 µg/kg and a limit of quantification (LOQ) of 2 µg/kg, ensuring reliable sensitivity (46). Microbiological analysis was performed according to the U.S. FDA *Bacteriological Analytical Manual* (BAM) protocol (47) (detailed in supplementary file).

### 2.3 Gamma Irradiation

Germinated (48 h) and roasted finger millet flour was irradiated with a minimum radiation dose of 8 kGy for 240 minutes and 18 seconds at the gamma irradiation unit of Gamma Agro Medical Processings Pvt. Ltd, Hyderabad, Telangana, in accordance with ISO 11137-1 guidelines (48). Food irradiation practices are also supported by FAO/IAEA/WHO recommendations, which recognize doses up to 10 kGy as safe for improving microbiological safety and extending shelf life (49), (50), (51), (52), (53), (54), (55). This gamma-irradiated finger millet flour was then used in the preparation of FMCF.

### 2.4. Sensory Evaluation and Acceptability Study

Sensory evaluation was conducted at the ICMR–National Institute of Nutrition (NIN) and nearby Anganwadi Centres during morning hours to assess product characteristics and acceptability. Based on repeated-measures ANOVA (3 products, 5-point hedonic scale, α = 0.05, power = 80%, effect size = 0.25), a sample size of 28 was estimated; hence, 30 untrained panelists (ICMR-NIN staff, students, Anganwadi teachers, caregivers, and mothers aged 20–49 years) were recruited. Participants without sensory or health impairments evaluated freshly prepared samples (10 g each) served in coded paper cups under uniform lighting. Using a 5-point Hedonic scale (5 = like extremely to 1 = dislike extremely), panelists rated seven attributes—appearance, colour, odor, flavour, texture, taste, and overall palatability—for two test products (F1 and F2) and a control. Samples were presented randomly, and palate cleansing with water was ensured between tastings.

A test-meal feeding trial involving 30 children under five years from selected Anganwadi Centres was conducted to assess product acceptability. Using a purposive sampling strategy, each child received one recipe daily for five days under caregiver supervision. A sample size of 30 was considered suitable for pilot feasibility testing (56). After a one-hour fasting period, pre-weighed servings (60–120 g) were provided, and consumption was measured by re-weighing feeding vessels post-meal; intake of ≥ 50% was deemed acceptable. Health status, feeding time, and tolerance were monitored throughout, and any signs of illness or discomfort were recorded. All the obtained data of nutrient analysis were evaluated as Mean and SD. Data on sensory attributes of developed products and recipes were subjected to One-way ANOVA using Kruskal-Wallis test and the Independent sample t-test to compare among samples.

### 2.5. Intervention Design and Trial Protocol

#### 2.5.1 Inclusion Criteria for MAM

Children aged 18 to 59 months (below 5 years) with primary Moderate Acute Malnutrition (MAM) were enrolled. Eligibility criteria included a weight-for-height z-score (WHZ) ranging from -2 to -3, following WHO child growth standards. Only individuals diagnosed with MAM for the first time and with no history of prior treatment for undernutrition, as verified through records at Anganwadi centres, were included.

#### 2.5.2 Exclusion Criteria for MAM

Children with severe anemia, defined as hemoglobin levels below 7 mg/dl (assessed by Horiba), were excluded. Additionally, those who have consumed any products significantly modify the gut microbiome (such as antibiotics, probiotics, prebiotics) within four weeks prior to enrolment, were excluded. Children with a recent history of diarrhea, severe acute respiratory illness (SARI), or other illness necessitating hospitalization were deemed ineligible. Furthermore, children with congenital or acquired disorders impacting growth, such as trisomy 21 or cerebral palsy, or those undergoing concurrent treatment for other medical conditions, were similarly excluded.

#### 2.5.3 Recruitment and Screening for Eligibility

Accurate anthropometric measurements are essential to determine whether a child falls into the MAM or SAM category. These measurements were conducted by expert field personnel from the ICMR-National Institute of Nutrition (NIN). The field team visited the designated Anganwadi Centres (AWCs) to carry out the screening, assisted by trained field and nursing staff. General information—including the child’s name, mother’s name, date of birth (verified using the birth card), birth weight, and chronic disease history—was collected using a structured general information sheet. Anthropometric measurements—including height, weight, and mid-upper arm circumference (MUAC)—were recorded using calibrated instruments (Seca scales). For children under 24 months, recumbent length was measured using an infantometer. Multiple readings were taken by trained staff to minimize inter- and intra-observer variability, ensuring data accuracy and reliability.

#### 2.5.4 Data collection and management

### Anthropometry and general information

Data were collected using the ‘EpiInfo App,’ developed by the CDC (Centre for Disease Control and Prevention) and locally adapted by NIN developers. This platform was consistently used across all five time points to capture anthropometric and general information. Data were compiled centrally and analyzed using WHO Anthro and other appropriate software. Classification of nutritional status was based on the WHO growth standards. Children identified with MAM were selected as study participants, and age- and gender-matched healthy controls were also enrolled. Eligibility was confirmed through a structured questionnaire-based interview conducted by trained study staff. Clinical examination of eligible children was performed by trained physicians using a standardized clinical assessment form.

### Diet data collection

Dietary assessment was performed using two methods: a three-day 24-hour dietary recall and a Food Frequency Questionnaire (FFQ). The recall captured all foods, beverages, and snacks consumed along with their timing, preparation methods, and ingredients. The FFQ captured the habitual dietary intake for an extended period, documenting frequently consumed foods, their frequency (daily, weekly, or monthly), and typical portion sizes. Portion sizes for both methods were measured using calibrated weighing scales and estimated using food models, standard serving sizes, or reference photographs. For home-cooked foods, raw ingredient amounts were derived using yield factors from the Indian Food Composition Tables (IFCT). Food items were grouped into culturally relevant categories such as cereals, pulses, fruits, vegetables, dairy, animal proteins, fats, and snacks. All dietary responses were digitized and entered into a central database for planned analyses, including calculating Nutrient adequacy, Diet-Diversity Score (DDS), etc.

#### 2.5.5 Enrolment

Children who met all screening and clinical eligibility criteria were enrolled as study participants and assigned a Unique Identification Number (UID) for tracking. Informed consent was obtained from caregivers (two languages), and a project information sheet was shared with AWTs. Mothers (or available guardian) were briefed on the purpose, procedure, estimated time, and probable usage of the results at the state/national level. Also, the member (s) of the family, who usually cook and serve the food (usually the home maker), was/were identified as our respondent.

#### 2.5.6 Training and Demonstration

Prior to the supplementation phase, the concerned caregivers/ AWTs were trained by the project team on individual serving sizes (based on child body weight), recipe preparation, measurement techniques, and hygiene practices. The training aimed to standardize food preparation and minimize errors during feeding.

#### 2.5.7 Sample collection and storage

##### Fecal samples

Baseline fecal samples were collected immediately prior to the feeding trial, following a detailed SOP. Parents were instructed on sample collection procedures and provided sanitized collection kits along with guidance on hygiene and sample handling. Collected samples were stored in dry ice containers, transported under cold chain conditions, and stored at –80°C until further analysis.

##### Blood samples

Blood samples were drawn by trained phlebotomists with minimal discomfort to the child. Samples were transported promptly to the laboratory, where they were processed to obtain serum and plasma, and stored at –80°C until further use.

#### 2.5.8 Supplementation

Following quality control, nutritional analysis, and sensory evaluation, the finger millet-based (FMD) and wheat-based (Bal-D) premixes were distributed to AWTs and caregivers. Calculated servings were administered to participants as per guidelines. During the first week of supplementation, a project team member was present at each centre to assist and ensure protocol adherence. To monitor compliance, food diary forms were provided to caregivers for daily updates on attendance and food consumption. Weekly supplies of premixes were delivered by the field team.

##### Regular visits

Throughout the intervention period, the project team made alternate-day visits to AWCs to oversee supplementation. Each centre was assigned a team member responsible for ensuring proper record-keeping and food intake monitoring. Daily food diaries were regularly reviewed, and a weekly ‘Adverse Events Form’ was used to document any health issues such as changes in appetite, vomiting, diarrhea, cough, cold, or fever.

##### Post-Trial Care

No specific post-trial care or intervention has been planned as part of this study. During the trial period, a pediatrician and clinicians has been made available to monitor and manage any adverse events promptly. After the trial concludes, no further clinical care or follow-up is required as the intervention does not necessitate continued administration or monitoring.

#### 2.5.9 Outcome measures Primary Outcome

The primary outcome measures were the efficacy of the developed millet-based food in improving nutritional status and its gut-modulatory effects in children with moderate acute malnutrition (MAM). Specifically, this included changes in anthropometric indicators such as weight-for-age and height-for-age z-scores, alongside modulation of gut microbiota composition and diversity assessed through metagenomic or 16S rRNA sequencing.

##### Secondary Outcome

Secondary outcome measures included quantification of gut inflammatory and nutritional biomarkers to evaluate the immunomodulatory effects of the intervention. Additionally, assessment of the long-term sustainability of nutritional improvements and microbial changes has been considered through follow-up measurements beyond the intervention period.

## 3. Sequencing and Analysis plan

### Microbial composition and inflammatory marker analysis

A 200mg stool sample will be taken for the DNA extraction. The extraction will be performed using the QIAamp PowerFecal Pro DNA Kit from Qiagen. The kit uses a novel and proprietary method for isolating both microbial and host genomic DNA from stool and gut samples. The kit uses QIAGEN’s second-generation Inhibitor Removal Technology (IRT), intended for use with samples containing inhibitory substances commonly found in stool, such as polysaccharides, heme compounds, and bile salts. The kit uses column-based separation of the DNA (silica membrane).

Microbiome compositional analysis will be performed using PacBio full-length long read sequencing approach, covering all the hypervariable regions (V1-V9) of 16S rRNA. This will be done utilizing single-molecule real-time (SMRT) sequencing and circular consensus sequencing (CCS), generating highly accurate long reads. Bacterial richness and abundance will be estimated for the samples, covering different timepoints of analysis.

Gut inflammation will be assessed using ELISA-based estimation of stable gut inflammatory molecules from fecal samples. Other nutritional markers and systemic inflammatory markers will be evaluated from blood using autoanalyzer instruments and ELISAs.

### Statistical plan

Associations between microbial profiles, gene expression, and clinical variables will be analyzed using multivariate methods. Tools like MaAsLin (huttenhower.sph.harvard.edu/galaxy) will employ boosted additive general linear models to assess relationships between predictors (metadata) and responses (microbial abundance or function), adjusting for age, gender, and other relevant factors influencing undernutrition phenotypes and microbial diversity.

Relative and differential bacterial abundance, along with alpha and beta diversity indices, will be visualized using tools like QIIME2 and DADA2. Group differences in gut microbiota composition at each time point will be analyzed with PERMANOVA on Bray-Curtis dissimilarity matrices and LEfSe to identify differentially abundant taxa. Changes in inflammatory markers will be evaluated via ANOVA, while their correlations with gut microbiota changes will be assessed using Spearman or Pearson correlation coefficients.

Multiple linear regression will be used to evaluate the associations between specific microbial taxa and inflammatory markers, while controlling for potential confounders.

## 4. Results

### 4.1. Nutrient Composition and Bioactive Results

Two products were developed based on the existing Balamrutham Plus (RUTF) formulation (Table 1). The Finger Millet Complementary Food (FMCF; F1) incorporated germinated-roasted finger millet and date powder, replacing 40 g of roasted wheat and rice flakes from the original formulation. In contrast, RUTF-2 (F2) substituted only sugar with date powder, retaining other ingredients. Common components across all formulations included skimmed milk powder, roasted Bengal gram, peanuts, and oil.

The total energy content of F1 and F2 was 437 kcal and 434 kcal per 100 g, respectively, providing more than 4 kcal/g of energy, in line with CODEX Alimentarius (2017) standards for therapeutic foods. Both contributed approximately 40–50% of daily energy requirements for under-five children (57), ICMR-NIN, 2020. The protein-energy ratio (PER) for F1 (10.54%) and F2 (10.47%) met WHO and CODEX recommendations (6–15%), with F1 showing a higher protein level due to the inclusion of germinated finger millet. The fat-energy ratio (FER) of 45.5% for both fell within the WHO-specified range (45–60% TE).

F1 exhibited the highest dietary fibre content, contributed by finger millet and date powder, and met the Indian recommended adequate intake (AI) for healthy children (15–20 g/day). F1 also demonstrated superior mineral composition, particularly calcium (461 mg), magnesium (171 mg), iron (3.1 mg), and zinc (3.1 mg) per 100 g, outperforming both F2 and the existing RUTF.

Bioactive analysis revealed significantly higher levels of total phenolic content (1118.43 ± 46.98 mg GAE), flavonoids (554.78 ± 17.58 mg CAE), and antioxidant activity (31.26%) in F1, followed by F2 and RUTF (p < 0.001). The inclusion of germinated-roasted finger millet and date powder substantially enhanced polyphenolic and antioxidant potential.

### 4.2. Sensory and Acceptability Outcomes

Sensory evaluation of the formulated mixes (F1 and F2) compared with the control (RUTF – Balamrutham Plus) showed high acceptability across all products. Using a 5-point Hedonic scale, no significant differences were observed for appearance, aroma, texture, taste, flavour, or overall acceptability (p > 0.05), except for colour, where F1 differed significantly from the control (p < 0.05). Overall acceptability scores ranged from 4.6 to 4.8, indicating good panelist response (refer Figure_4).

Evaluation of the four traditional recipes: laddu, porridge, idly, and pancake, showed recipe-specific variations. F1 laddu achieved the highest overall acceptability (4.8 ± 0.1), while F2 laddu scored lower for taste and acceptability (p < 0.05). F1 porridge outperformed the control in texture and consistency (p < 0.001) and was well accepted, whereas F2 porridge received lower scores for appearance, colour, and flavour. F1 idly recorded the lowest sensory scores (p ≤ 0.001), likely due to its darker colour and earthy taste, while F2 idly showed better scores (4.6 ± 0.1). Both F1 and F2 pancakes scored lower than the control (p < 0.05), though F2 pancake had better overall acceptability (4.2 ± 0.2). Overall, F1-based laddu and porridge and F2-based idly and pancakes were most preferred (Figure_5).

Children in both intervention groups: F1 (finger millet + dates) and F2 (Balamrutham + dates) showed significantly higher consumption of millet-based recipes compared to the control diet (p < 0.001, Table 2). Mean consumption ranged from 81–98% for F1 and 83–91% for F2, while the control group recorded only 53–71%. Feeding durations were generally shorter for millet-based recipes, indicating greater ease of intake and palatability. Among the recipes, F1 porridge and pancake achieved the highest acceptance (>95% consumption) and the shortest feeding times, while F2 idly, though well accepted, required a longer feeding duration.

### 4.3. Product Safety and Shelf-Life Validation

No aflatoxin contamination was detected in either the germinated-roasted finger millet or roasted peanut samples, confirming the safety of raw ingredients. Microbial analysis conducted on raw, germinated-roasted, and gamma-irradiated finger millet flours, as well as the final formulations (F1 and F2), showed a substantial reduction in microbial load after processing. The raw flour exhibited a high total colony count, which decreased markedly after germination and roasting, indicating the efficiency of heat treatment and low moisture in microbial reduction. E. coli and Salmonella were absent in all samples. Following gamma irradiation, no microbial colonies were detected in the germinated-roasted sample (GRFM-48h), confirming complete microbial elimination. Both F1 and F2 complied with FSSAI (2011) microbiological safety standards. Shelf-life testing using the USFDA-BAM protocol indicated an increase in total plate count (TPC) on day 30; however, initial values 1.2 × 10³ CFU/g for FMD and 8.0 × 10³ CFU/g for BMD were well within acceptable limits.

## 5. Discussion

The improved nutritional and functional profile of FMCF (F1) highlights the synergistic benefits of incorporating germinated-roasted finger millet and date powder. Germination enhanced nutrient bioavailability, amylase activity, and energy density while reducing bulk, as recommended by ICMR-NIN dietary guidelines. Roasting improved flavour, lowered antinutrient content, and enhanced palatability. Compared with the existing wheat–rice-based RUTF, F1 provided higher protein, fibre, and mineral levels, particularly calcium and iron, crucial for bone health and growth in malnourished children.

The elevated polyphenol and flavonoid contents in F1 contributed to greater antioxidant capacity, supporting oxidative stress reduction and gut health restoration. Previous studies corroborate these findings, reporting enhanced bioaccessible polyphenols after germination (20–67% increase) and roasting (17% increase) (43). Overall, F1 emerges as a nutrient-dense, microbiota-supportive complementary food, suitable for MAM management, while F2 provides a healthier sugar-free alternative aligned with long-term child health goals.

The absence of aflatoxin contamination and the reduced microbial load confirm that roasting, germination, and gamma irradiation are effective in ensuring product safety. These findings align with previous reports highlighting the role of heat and irradiation in lowering fungal and bacterial contamination. The slight increase in microbial counts during storage, similar to trends in other nutrient-rich RUTFs (59), remained within FSSAI safety limits, indicating that both F1 and F2 are microbiologically stable and safe for consumption.

Sensory results demonstrate that both formulations were well accepted, with sensory characteristics comparable to the control, confirming the feasibility of incorporating germinated finger millet and date powder in complementary food formulations. The high acceptability of F1 laddu and porridge can be attributed to the natural sweetness and binding quality of date powder and the smooth texture imparted by germinated finger millet, consistent with earlier findings by (60). Although F1 idly and pancakes scored lower due to their darker colour and earthy flavour, F2 recipes achieved better sensory appeal, likely due to their lighter colour and milder taste. These outcomes align with previous studies reporting high sensory acceptance of millet-based complementary foods among children and adults (61) . Overall, the results confirm that finger millet–based formulations can be successfully used in traditional recipes without compromising consumer acceptance.

The high consumption rates and reduced feeding times of millet-based recipes indicate superior palatability, texture, and ease of swallowing, especially in F1 preparations. These findings are consistent with earlier reports showing >90% consumption of finger millet porridge by children in community settings (62). The improved acceptability of F1 and F2 over the control highlights the role of germinated finger millet and date powder in enhancing flavour, sweetness, and texture. Similar trends have been reported where millet incorporation in traditional foods improved both sensory appeal and nutrient intake (63), (64). Overall, the results support the feasibility of integrating millet-based formulations into child feeding programs as culturally acceptable and nutritionally superior alternatives to conventional cereal-based diets.

## 6. Expected Outcomes and Impact

This community-based randomized trial is designed to advance the field of pediatric nutrition. Most existing interventions primarily target complicated forms of undernutrition, while uncomplicated states often remain overlooked. Conventional calorie-dense approaches focus on energy repletion but fail to address the underlying biological dysfunctions. As a result, recovery is often short-lived, and relapse into malnutrition is common. This study seeks to bridge the gap between nutritional recovery and gut ecological restoration. It aims to evaluate the efficacy of finger millet-based nutritional and prebiotic supplements on the overall nutritional and gut health of children under five years of age with moderate acute malnutrition (MAM). Undernourished children frequently present with an inflamed gut environment. Inflammation draws excess oxygen into the lumen, disrupting normal anaerobic conditions and impairing host–microbe interactions. This results in poor nutrient assimilation and further perpetuates the cycle of undernutrition. The proposed supplement is expected to provide dual benefits: nutritional support and modulation of gut microbial composition. Finger millet has unique biological potentials that make it suitable for this purpose. They may contribute to ensuring long-term advantages by mitigating inflammation and preventing relapse following the completion of the intervention. The cultural acceptance and widespread use of finger millet also make it a practical option for integration into existing public health programs such as the Take-Home Ration scheme under ICDS, aligning with SDGs 2 and 3.. By generating robust clinical evidence, this study aims to establish microbiota-directed complementary feeding as a transformative strategy in managing childhood undernutrition. The focus thus shifts from merely addressing caloric needs to restoring resilient, metabolically active guts that can support sustainable child health and development.

## 7. Conclusions

The Finger Millet–Based Complementary Food (FMCF) was developed using a carefully optimized process involving 24-hour soaking, 48-hour germination, and roasting, which maximized nutrient retention and minimized antinutritional factors. This sequence produced a formulation rich in calcium, magnesium, fibre, and antioxidants, with enhanced mineral bioavailability. Incorporating date powder as a natural sweetener further improved the nutrient profile by adding fibre, iron, and potassium while avoiding refined sugar. Recipes prepared with FMCF demonstrated higher acceptability among children than the existing RUTF, highlighting its potential as a nutrient-dense, culturally acceptable, and sustainable intervention to address malnutrition and hidden hunger in young children.

## 8. CRediT authorship contribution statement

Authorship contributions: Sakshi Rai: Investigation, Methodology, Writing original draft; Nikhita BR: Investigation, Methodology, Writing original draft; Sahil Bipin Kumar Suthar: Formal analysis, Writing-Review and editing; Parth Sarin: Writing-Review and editing, software, data curation; Akshay Shinde: Data curation, Formal analysis; Aruna V. Reddy: Data collection, curation and Formal analysis; Vijayalakshmi G: Data collection, curation and Formal analysis; Santosh Kumar Banjara: Supervision, Validation, Visualization, Writing-Review and editing; Hemant Mahajan: Software, Supervision, Validation, Visualization, Writing-Review and editing; Ananthan Rajendran: Conceptualization, Methodology, Supervision, Validation, Visualization, Writing-Review and editing; Naveen Kumar R : Methodology, Supervision, Validation, Visualization, Writing- Review and editing; Radhika M S: Methodology, Supervision, Writing-Review and editing; Sourav Sengupta: Conceptualization, Funding acquisition, Investigation, Project administration, Supervision, Visualization, Writing-Review and editing; Devaraj J. Parasannanavar: Conceptualization, Funding acquisition, Investigation, Project administration, Supervision, Visualization, Writing-Review and editing.

## 10. Institutional Review Board Statement

Approved by the Institutional Ethics Committee (IEC No. - CR/2/V/2023); registered with Clinical Trials Registry—India (CTRI No. CTRI/2023/06/053590).

## 11. Informed Consent Statement

Informed consent was obtained from the parents/guardians of all the participants involved in the study. The consent form was provided in two languages: one local and one formal (Telugu and English).

## 12. Data Availability Statement

The data supporting this study’s findings will be made available on reasonable request.

## 14. Confidentiality and Dissemination policy

Participant data will be kept confidential and securely stored, accessible only to authorized personnel. Identifiable information will be coded to protect privacy. Results will be shared through publications and presentations without revealing participant identities, following ethical and regulatory standards.

## Supplementary Material

