## Supplemental File 1 for "Acceptability and gut-modulatory effects of Finger millet-based complementary food (FMCF) in treating Indian children with Moderate Acute Malnutrition: A randomized controlled trial protocol"

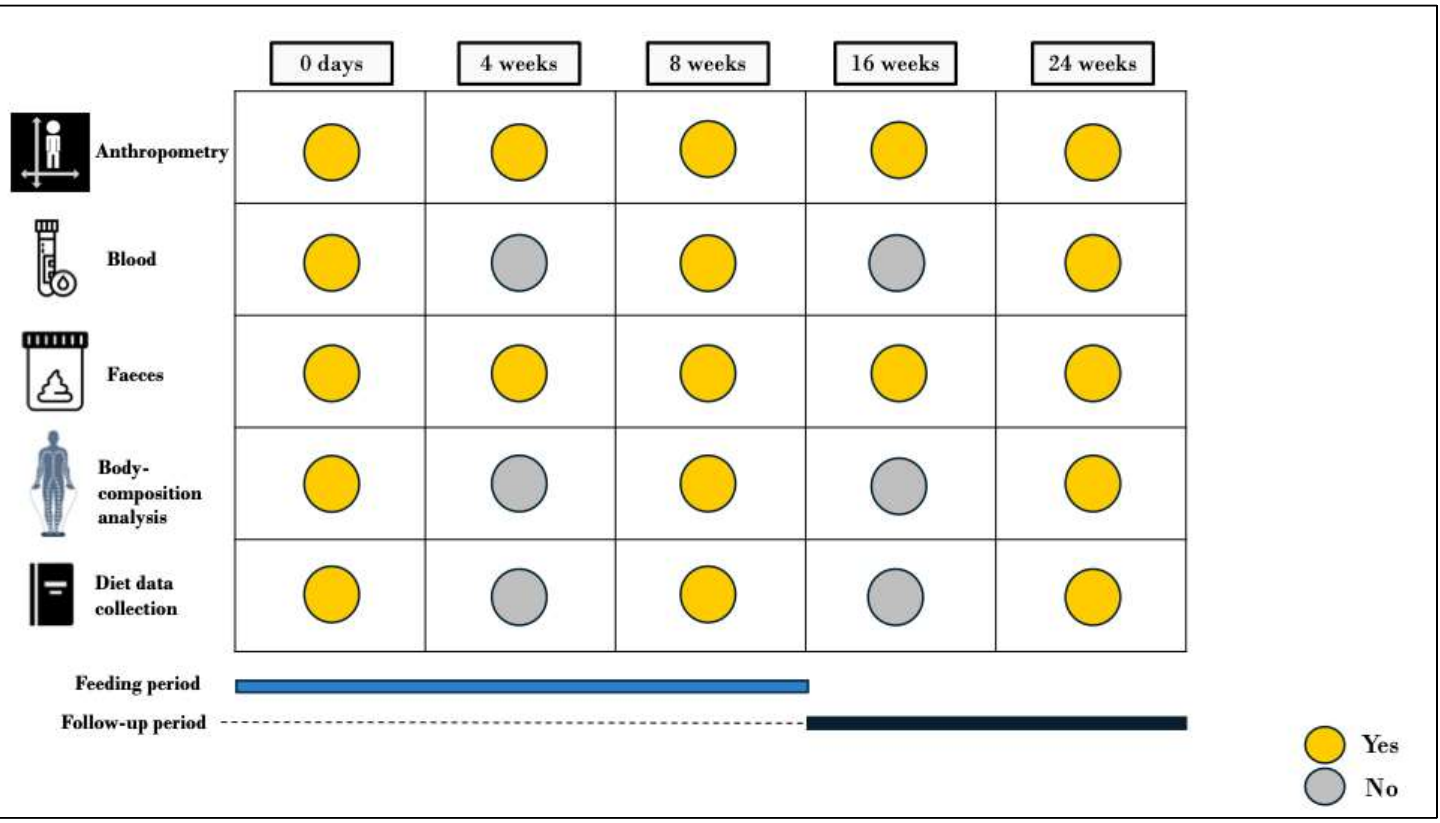

Figure_1: Study timeline showing schedule of assessments during the feeding and follow-up periods.

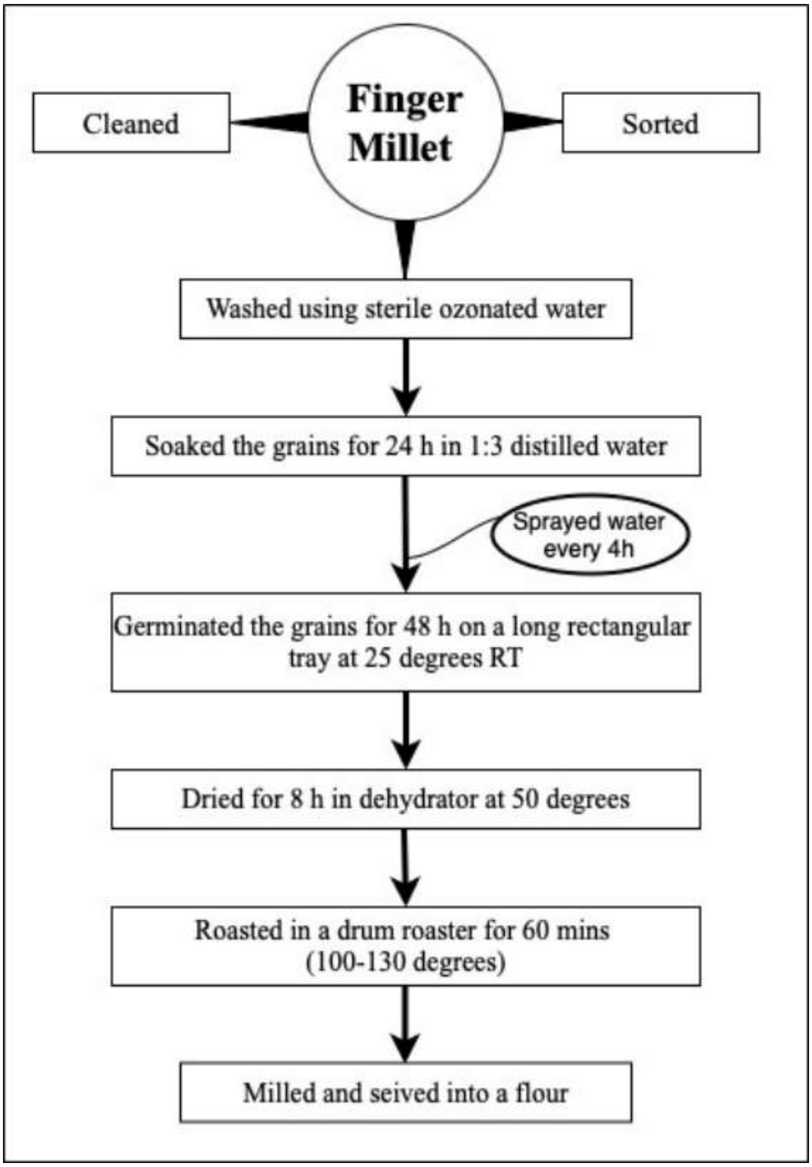

Figure_2: A flow chart diagram depicting the finger millet processing

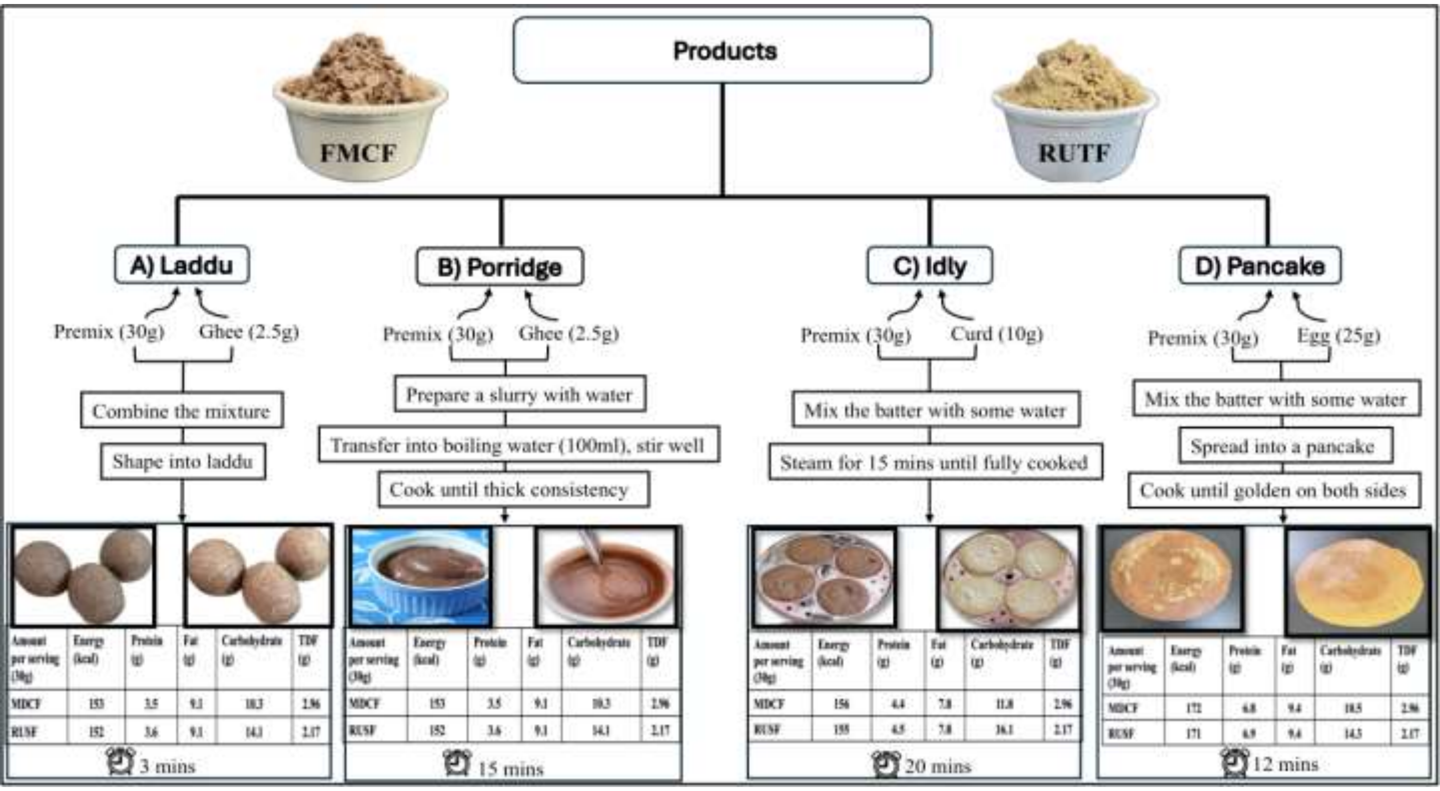

Figure_3: Flowchart of the developed four recipes using products (premix) with their nutritional information.

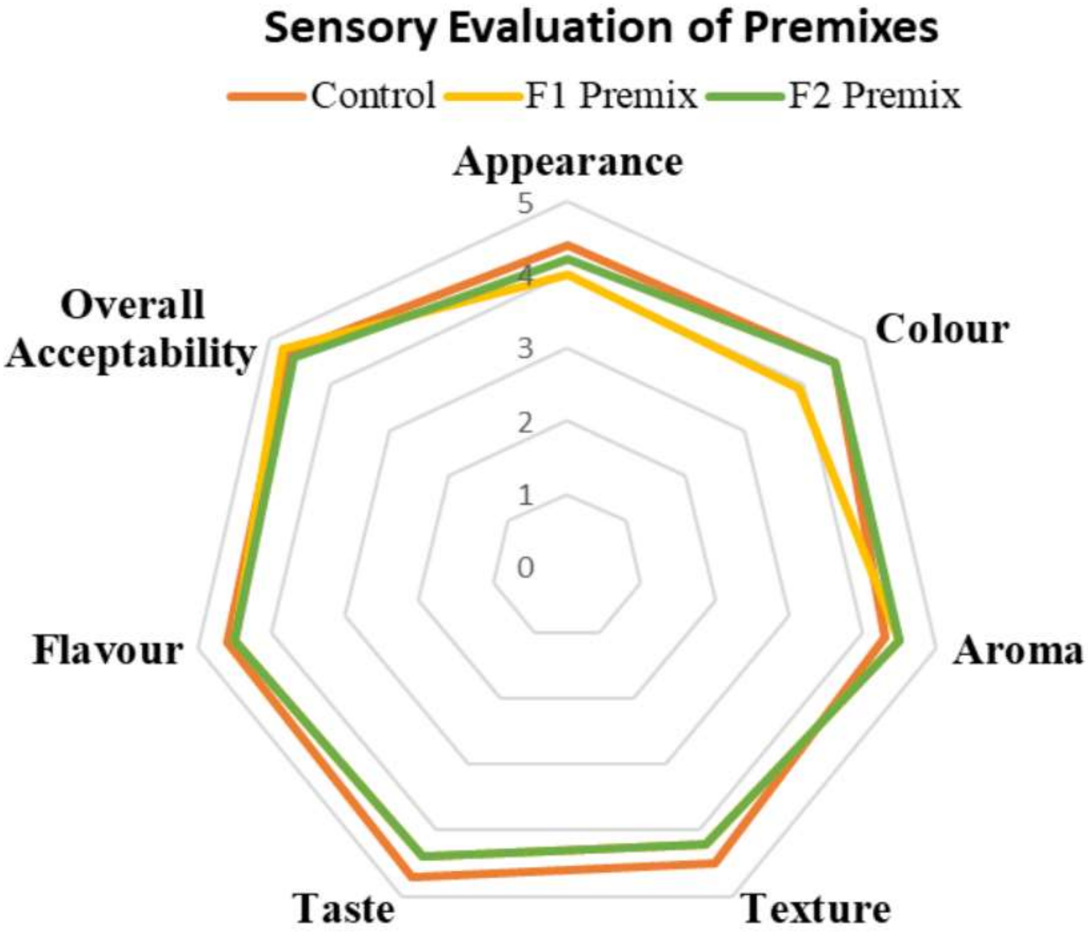

Figure_4 : Plot represents acceptability outcomes for the three tried premixes (F1, F2 and Control).

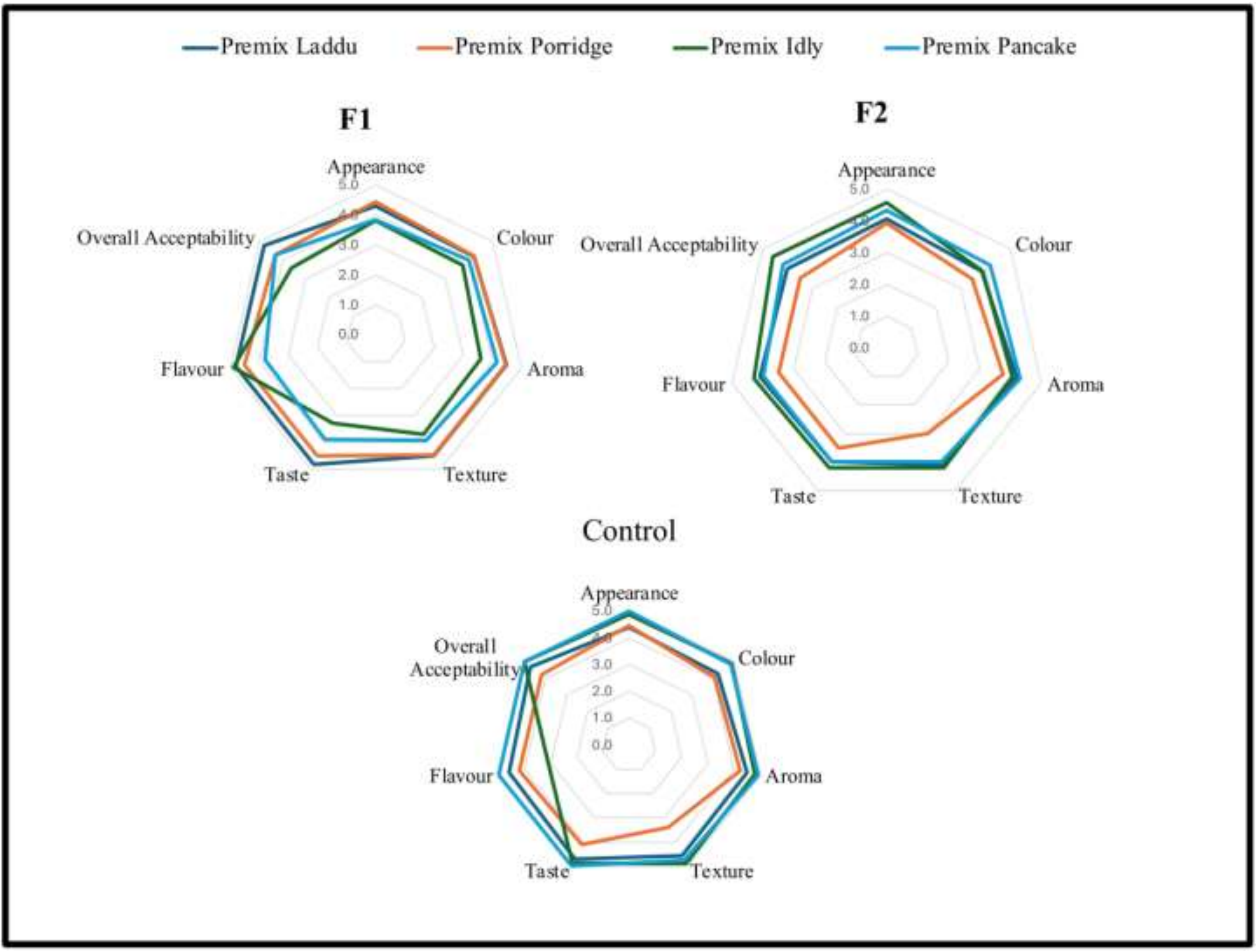

Figure_5: Plots represent the acceptability scores for the different tested recipes of F1, F2 and Control.

### Sensory Evaluation Form for the Institutional Sensory Panel

**Project title: Effect of finger millet based dietary supplementation on gut microbiota composition and function in uncomplicated moderate acute malnutrition in under 5 children**

**1. Participant ID: __ __ __ __ 2. Name of the Division: ___________________**

**3. Age (Years): __ __ 4. Gender (1. Male 2. Female): _________**

**5. Date of evaluation (dd/mm/yy): __ __/__ __ /__ __ 6. Recipe Code: _____________**

**7. Start Time (hh/mm): __ __ / __ __ 8. End Time (hh/mm): __ __ / __ __**

The Dietary Supplementation of Finger millet-based product is proposed for children in the age group of 24-54 months to satisfy the nutritional requirements and for the treatment of uncomplicated MAM. Please assess the sensory properties of the foods form left to right only, and give appropriate scores. Also, give suggestions if any, for improvement.

| SI NO | Sensory Attribute | Food Product Code<br>— — — | Food Product Code<br>— — — | Food Product Code<br>— — — | Comments / Suggestions |
|---|---|---|---|---|---|
| 01 | Appearance | | | | |
| 02 | Colour | | | | |
| 03 | Aroma | | | | |
| 04 | Texture/Consistency | | | | |
| 05 | Taste | | | | |
| 06 | Flavour | | | | |
| 07 | Overall Acceptability | | | | |

### Score: 5) Like extremely; 4) Like moderately; 3) Neither like nor dislike; 2) Dislike moderately and 1) Dislike extremely

Signature of the Evaluator

Signature of the Investigator

### Sensory Evaluation Form for the Institutional Sensory Panel

**Project title: Effect of finger millet based dietary supplementation on gut microbiota composition and function in uncomplicated moderate acute malnutrition in under 5 children**

**1. Participant ID: __ __ __ __ 2. Name of the Division: ___________________**

**3. Age (Years): __ __ 4. Gender (1. Male 2. Female): _________**

**5. Date of evaluation (dd/mm/yy): __ __/__ __ /__ __ 6. Recipe Code: _____________**

**7. Start Time (hh/mm): __ __ / __ __ 8. End Time (hh/mm): __ __ / __ __**

The Dietary Supplementation of Finger millet-based product is proposed for children in the age group of 24-54 months to satisfy the nutritional requirements and for the treatment of uncomplicated MAM. Please assess the sensory properties of the foods form left to right only, and give appropriate scores. Also, give suggestions if any, for improvement.

| **SI NO** | **Sensory Attribute** | **Food Product Code** ___ ___ ___ | **Food Product Code** ___ ___ ___ | **Food Product Code** ___ ___ ___ | **Comments / Suggestions** |
|---|---|---|---|---|---|
| 01 | Appearance | | | | |
| 02 | Colour | | | | |
| 03 | Aroma | | | | |
| 04 | Texture/Consistency | | | | |
| 05 | Taste | | | | |
| 06 | Flavour | | | | |
| 07 | Overall Acceptability | | | | |

### Score: 5) Like extremely; 4) Like moderately; 3) Neither like nor dislike; 2) Dislike moderately and 1) Dislike extremely

Signature of the Evaluator

Signature of the Investigato

### Test-Meal Feeding Trial Study

**Project title: Effect of finger millet based dietary supplementation on gut microbiota composition and function in uncomplicated moderate acute malnutrition in under 5 children**

**Date: __ __/__ __/__ __**

a) **Anganwadi Centre Name: ________________________________**
b) **Recipe Code: _ _ _**
c) **Name of the Recipe: ___________________**
d) **Product Code: _ _ _**

| S.No. | Name of the subject | Gender | Age (Yrs) | Weight (Kg) | Meal time | Serving size (g) | Leftover (g) | Consumption (%) |
|---|---|---|---|---|---|---|---|---|
| 1 | | | | | | | | |
| 2 | | | | | | | | |
| 3 | | | | | | | | |
| 4 | | | | | | | | |
| 5 | | | | | | | | |
| 6 | | | | | | | | |
| 7 | | | | | | | | |
| 8 | | | | | | | | |
| 9 | | | | | | | | |
| 10 | | | | | | | | |
| 11 | | | | | | | | |
| 12 | | | | | | | | |
| 13 | | | | | | | | |
| 14 | | | | | | | | |
| 15 | | | | | | | | |
| 16 | | | | | | | | |
| 17 | | | | | | | | |
| 18 | | | | | | | | |
| 19 | | | | | | | | |
| 20 | | | | | | | | |

**Signature of the Evaluator** **Signature of the Investigator**

#### Screening Form

| | | |
|---|---|---|
| Anganwadi Location | | |
| Anganwadi Teacher Name | | |
| Anganwadi Contact Number | | |
| Date of Subject Enrollment | | |
| Subject ID | | |
| Subject Name | | |
| Age | DOB: | |
| Sex | Male | Female |
| Weight (kg) | | |
| Height (cm) | | |
| MUAC (cm) | | |
| Z- Score | | |
| Birth weight | | |
| Father's/Mother's Name | | |
| Contact Number | | |
| Any history of illness in past 4 weeks (Fever/Cold/Cough) | Yes | No |
| If yes (Any medication) | | |

| CLINICAL EVALUATION | | | | | |
|---|---|---|---|---|---|
| **QUESTIONS** | **ANSWERS option** | | | | |
| Allergy information | Yes | No | If yes, enter manually | | |
| Food allergy | Yes | No | If yes, enter manually | | |
| Intolerance (if any) | | | | | |
| | Milk intolerance | | | | |
| | Millet intolerance | Other, mention | | | |
| Did the child attended last deworming schedule (within 6 months) | Yes | No | | | |
| Behavioural disorder | Apathy | Irritability | Impaired social ineraction | Other | |
| Eye examination | Normal | Pale | Spots | Vitrous | Other, mention |
| Skin appearance | | | | | |
| Nail appearance | | | | | |
| Hair appearance | | | | | |
| Pica disorder | Yes | No | | | |
| Appetite | Poor | Good | Variable | | |
| Eating disorder | Yes (If yes add manually) | No | | | |
| Number of times of defecation per day | 1 to 2 times | > 2 times | once in two days | | |
| History of abdominal pain | | | | | |
| **DELAYED MILESTONE** | | | | | |
| | **Gross-motor development** | | **(Answers to be ticked)** | | |
| 2 years | Physical Activity of child- e.g., Playing | | | | |
| 3 years | Is the child able to pedal a bicycle | | | | |
| 4 years | Is the child able to hop | | | | |
| 5 years | Is the child able to skip | | | | |
| | **Fine-motor development** | | | | |
| 2 years | Is the child able to draw a line/ scribble | | | | |

| | | | |
|---|---|---|---|
| 3 years | Is the child able to draw a circle | | |
| 4 years | Is the child able to draw a square/ cross | | |
| 5 years | Is the child able to draw a triangle | | |
| | **Social/communication** | | |
| 2 years | Is the child able to talk in 2 word language | | |
| 3 years | Is the child able to talk in 3 word language | | |
| 4 years | Is the language 100% understandable by stranger | | |
| 5 years | Is the child able to talk in 5 word language | | |
| | **Cognitive/adaptive** | | |
| 2 years | Is the child able remove one of the clothing | | |
| 3 years | Is the child able to brush teeth with help | | |
| 4 years | Does he/she know 4 colours atleast | | |
| 5 years | Is he/she able to dress up on own | | |
| Antibiotic consumed within past 4 weeks | Yes | No | |
| If yes (Mention name) | | | |
| Probiotics consumed within past 4 weeks | Yes | No | |
| If yes (Mention name) | | | |
| Any other medication taken within past 4 weeks | Yes | No | |
| If yes (Mention name) | | | |
| History of seizures | Yes | | No |
| If yes (Type of seizures) | Febrile | | |
| | Afebrile | | |
| Has the child recently suffered fromPneumonia | Yes | No | |
| If Yes (Date) | | | |

| | | |
|---|---|---|
| Recent history of Jaundice | Yes | No |
| If yes (Date of diagnosis-referring report) | | |

**ICMR-NATIONAL INSTITUTE OF NUTRITION**

**JAMAI-OSMANIA (P.O.) HYDERABAD-500 007**

#### Subject Informed consent form

**"Effect of finger millet based dietary supplementation on gut microbiota composition and function in uncomplicated moderate acute malnutrition in under 5 children" .**

Center name :

Subject Full Name with Initials: ……………………………

Name of Father :

Name of Mother :

Age : …………………………....

I confirm that I have read and understood the information sheet about the above research study and have the opportunity to ask questions. I understand that the participation in the study is voluntary and I am free to withdraw at any time without giving reasons without my medical care or legal rights being affected.

**I understand that if my child is diagnosed with 'primary MAM' (and I agree to participate in this study), 5/6 ml of Venous Blood sample (thrice: Day 0, Day 30, Day 60) and one table spoon of fresh fecal sample** of my daughter /son will be collected **(five times: Day 0, Day 30, Day 60, Day 90, Day 120)** using all aseptic precautions with sterile containers/devices, for laboratory investigation. My daughter/son will be supplemented with finger millet/ wheat based dietary supplement.

OR, **I understand that if my child is diagnosed as 'Healthy' based on the study parameters (and I agree to participate in this study), one table spoon of fresh fecal sample** of my daughter /son will be collected **(three times: Day 0, Day 30, Day 60)** using all aseptic precautions with sterile containers/devices, for laboratory investigation. My daughter/son will be supplemented with wheat based dietary supplement containing either sugar or dates powder.

We are informed that taking those supplementation does not have any risk. I have been informed that confidentiality will be maintained about all my information.

I agree to participate in the above study.

**Signature (or thumb impression) of Parent/ Guardian: …………………………….**

**Date : …………………………………..**

**Study Investigator's Name : Dr. Devraj J.P.**
**Cell Number : 8074928620**
**Signature of investigator : ………………………………….**
**Signature of Witness :…………………… Name of witness :…………………………**

**Subject Information Sheet**

**"Effect of finger millet based dietary supplementation on gut microbiota composition and function in uncomplicated moderate acute malnutrition in under 5 children".**

**Subject recruitment and sample collection:**
**Laboratory testing:** ICMR- National Institute of Nutrition, Hyderabad
**PI : Dr.Devraj JP**
**Co-PI: Dr.Sourav Sen Gupta**
**Physicians: Dr.Santosh Kumar/ Dr.Samarsimha Reddy/Dr.Mahesh Kumar M**
**Date of Ethics Committee Approval:**
**-----------------------------------------------------------------------------------------------------------**

**Introduction:** I, **Dr. Devraj JP**, Scientist C' at National Institute of Nutrition, Hyderabad, requesting you to take part in a research study, which will let you know about "**Effect of finger millet based dietary supplementation on gut microbiota composition and function in uncomplicated moderate acute malnutrition in under 5 children**". Please read carefully the following explanation about this study and take as much time as you need to enroll your kid or not. If you do not understand or would like more information, please feel free to ask the investigator at any point of time**.**

**Purpose of the study:** Malnutrition especially undernutrition remains one of the most pressing global health challenges today, contributing to nearly half of all deaths in children under five years of age. In addition to insufficient dietary intake and environmental enteric dysfunction, gut microbiome impairment in these children is coming to light as a possible important contributor towards persistent growth impairment. Studies on the gut microbiome of children diagnosed with severe acute malnutrition (SAM) using metagenomic techniques have revealed drastic alterations in the microbiome structure and function and provided insights as to how these alterations could lead to growth impairment in children. Dysbiosis in the gut microbiota composition are now being causally linked to undernutrition. An altered gut microbiota in undernutrition often accompanied by low threshold of enteropathogens from an unsanitary environment triggers a subclinical constellation of intestinal pathologies that include inflammation.

We would like to conduct randomized control trials to test the effect of a finger millet based dietary intervention on the gut microbiome and its impact on the nutritional status measured by anthropometry of undernourished children from Anganwadi schools at Hyderabad.

**Your participation in this study and eligibility:**

Your kid must be 18-59 months who are diagnosed with primary moderate acute malnutrition (MAM: between -3 and -2 SD of WHO growth standards for median weight for height (wasting) z-scores respectively) and is presently in an uncomplicated state of health. Controls group will be defined as children aged 18-59 months from the same AWC who are not diagnosed with any grade of malnutrition. Subjects with MAM will be given 50gm of Finger millet based or Wheat based nutritional intervention for 8 weeks and followed up till 6 months. Controls will be provided with 50gm of either regular Balamrutham+ or modified Balamrutham+ nutritional intervention for 8 weeks. There will be no follow-up and no blood sample will be collected from child diagnosed as 'Healthy'. Fecal sample of the controls will be collected for 3 time points (0 day, 30$^{th}$ day, 60$^{th}$ day) for gut microbiota analysis.

You should be willing to give 5/6 ml of blood (thrice) and fresh fecal sample (five times) from your child during the study period if diagnosed as MAM. We will be profiling gut microbiota using the fecal samples. We will be doing anthropometry measurements, Clinical examination and detailed history concerned with health will be collected. If you are fitting the inclusion criteria and if you agree to participate in the study, then you will receive finger millet or wheat based dietary intervention and its impact on the gut microbiome and on the nutritional status will be measured by anthropometry after 8 weeks supplementation.

**Who is responsible for this study?**

**P.I: Dr.Devraj JP** Scientist C' Division of Clinical Epidemiology, ICMR- National Institute of Nutrition, Hyderabad, are responsible.

**Risks and Benefits of Participating in the study:**

**Benefits: Uncomplicated Primary MAM children** will be supplemented with finger millet/wheat based dietary intervention. Immediate benefit will be improvement in their nutritional health status and gut health.

**Risks:** There will be no risk involved in this study. There will be minimal discomfort in collecting Blood and fecal samples.

**Voluntary participation:**

Taking part in this study is voluntary. You will not lose any of your legal or ethical rights. You may withdraw from the study at any time and you are not obliged to give reasons.

**Confidentiality of Information Obtained:**

All the information collected from you will be written down by the investigator and will be kept safely at ICMR-NIN. The written material will not bear your name. It will have an identification number and if we need more information from you, we will get back to you with the help of this identification number. All the information gathered from you would be accessible only to the Principal Investigators.

**Compensation for Your Participation:** No compensation will be paid during the study.

**Adverse events:** We do not expect any adverse events due to the study procedures. However, in case of any adverse effects during the course of the study, you will be advised on further measures**.**

**If you need any information at any time during the course of the study you may reach the Investigator at the telephone no.**

**Dr. Devraj J.P.: 8074928620 or 040-27197215**

**Consent of the participant for the study:**

I have read this Information Form completely / this Information Form has been read out to me in my mother-tongue. All my doubts are cleared. I can withdraw participation from the study at any time. I have received and understood the information about my rights and guarantee about maintaining confidentiality about my personal information.

**I want to participate in this study myself by my own free will.**

**Ethics Committee Stamp of Certification** **Date of Expiry of Consent Validity**

**Please do not sign after date of expiration**

**Date:** **Name:**

**Signature**

**Effect of finger millet based dietary supplementation on gut microbiota composition and function in uncomplicated moderate acute malnutrition in under 5 children**

#### 1. Basic Information

**Date:________**

**UID: FM-____________**

**Collected By:**

- **Child's Name: ___________________________________________**
- **Child's Age: ___________________________________________**

#### 2. Parent/Guardian Information

#### Mother Information

- **Name: __________________________________**
- **Age : __________________________________**
- **DOB(dd/mm/yyyy): ___________________________________**
- **Occupation:_______________________________**
- **Education:___________________________________**

**History of any major ailments:**

☐Diabetes

☐Hypertension

☐Heart disease

☐Dyslipidaemia

☐Liver Disease

☐Thyroid

☐None

Other:____________________________________

**Phone Number:________________________________**

**Diet preference:**

☐Veg

☐Non-Veg

☐Veg + Egg

Height(CM):___________________

Weight(Kg):___________________

#### Father Information

- **Name: __________________________________**
- **Age : __________________________________**
- **DOB(dd/mm/yyyy): ___________________________________**
- **Occupation:_______________________________**
- **Education:___________________________________**

- **History of any major ailments:**
  ☐Diabetes
  ☐Hypertension
  ☐Heart disease
  ☐Dyslipidaemia
  ☐Liver Disease
  ☐Thyroid
  ☐None

  Other:____________________________________
- **Phone Number:________________________________**
- **Diet preference:**
  ☐Veg
  ☐Non-Veg
  ☐Veg + Egg

#### 3. Family Level details

- **Religion:**
  ☐Hindu
  ☐Muslim
  ☐Christian
  ☐Others
- **Caste:**
  ☐General/UR
  ☐OBC
  ☐SC
  ☐ST
- **Total number of people living in the household: ________________**
- **Education status of the head of the family:______________________**
- **Where is the cooking done:**
  ☐Inside
  ☐Outside
  ☐Separate Room
- **Do you have a separate room for kitchen:**
  ☐YES
  ☐NO

- **Drinking Water:**
  ☐Tap water
  ☐RO water

- **How many rooms in the household are used for sleeping:__________**
- **Is Toilet facility available in the household:**
  ☐YES
  ☐NO
- **Does this household have a BPL card:**
  ☐YES
  ☐NO

- **Total family income:__________________**

- **Does your household have the following:**?
  ☐Electricity
  ☐Matress
  ☐Pressure cooker
  ☐Chair
  ☐Cot/Bed
  ☐TV
  ☐Sewing machine
  ☐Mobile
  ☐Landline/telephone
  ☐Computer
  ☐Refrigerator
  ☐AC/Cooler
  ☐Washing machine
  ☐Watch/clock
  ☐Motorcycle/Scooter
  ☐Animal drawn cart
  ☐Car
  ☐Water Pump
  ☐Thresher
  ☐Tractor
  ☐Bicycle
  ☐Internet
  ☐Table
  ☐Electric fan

#### 4. Child General and Birth Details:

- **Twins?**
  ☐YES
  ☐NO
- **Birth Order:__________**
- **Number of Siblings:__________**
- **Number of children under 5 years:______________**
- **Gestational Age:**
  ☐Full term
  ☐Pre-term
- **Place of delivery:**
  ☐Home
  ☐Hospital
  ☐Others

  **Place of delivery(others):__________________________**

- **Mode of Delivery:**
  ☐Normal
  ☐C-section
  ☐Assisted- Vacuum /Forceps
  ☐Non-Assisted

- **Gestational Diabetes:**
  ☐YES
  ☐NO

- **Mother's weight at pregnancy(Kg):__________**

- **Birth weight of children(#.##)(Kg):____________**

- **Colostrum Intake:**
  ☐YES
  ☐NO

- **At what time feeding was initiated?**
  ☐Less than 1 hrs
  ☐Less than 6 hrs
  ☐Within a Day

- **Exclusively breast fed (in months):____________**

- **Age of introduction of complimentary food(in months):___________**

- **Type of complementary food:**
  ☐Wheat based
  ☐Rice based
  ☐Both Rice and Wheat based
  ☐Others

Others specify:____________________

- Is the child still breast fed right now?
  ☐YES
  ☐NO

- Washing of child's hand with soap after toilet?
  ☐YES
  ☐NO

- Number of times breast-fed per day?: ________________

- **Diet preference:**
  ☐Veg
  ☐Non-Veg
  ☐Veg + Egg

- Has the enrolled child taken deworming medication in 2024:
  ☐YES
  ☐NO

#### 5. Other Information

- Screen time(Minutes):__________________
- Play time(Minutes): ____________________

### Recipe Manual

- **Recipe No.1: Laddu**

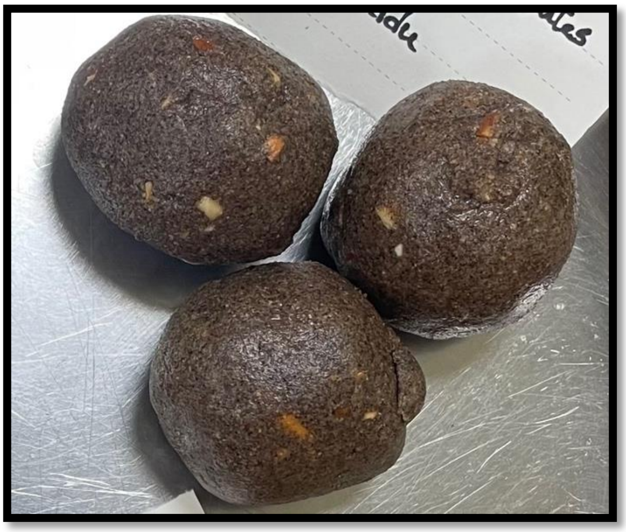

| **Ingredients** | **Quantity (g)** |
|---|---|
| Premix powder | 30 |
| Ghee | 2.5 (1 tsp) |
| Elaichi powder | A pinch |

**Preparation time**: 2-3mins

**Method**:

i. Weigh 30g of premix flour using weighing scale or scoop (30g scoop size).
ii. Add 2.5g ghee into the preheated pan and allow it to melt.
iii. Transfer the ghee to the weighed premix in a vessel.
iv. Further add a pinch of (elaichi) cardamom powder and mix it.
v. Gently mix until it attains a laddu form.and ready to serve.

- **Recipe No.2: Porridge**

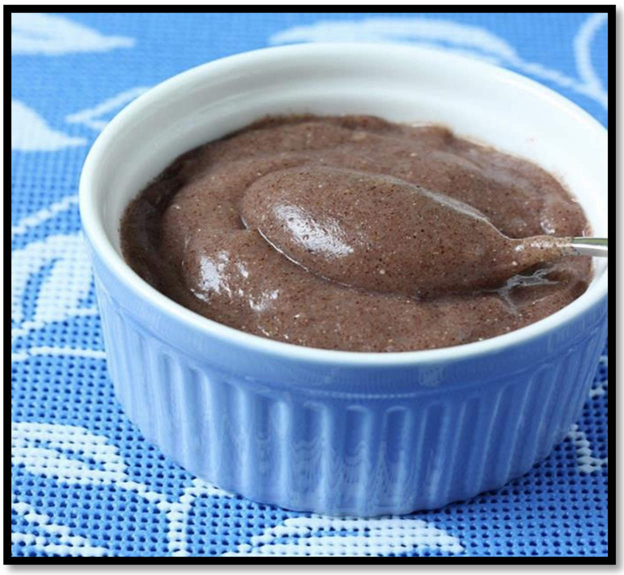

| **Ingredients** | **Quantity (g)** |
|---|---|
| **Premix powder** | **30** |
| **Ghee** | **2.5 (1 tsp)** |
| **Water** | **200ml (1 glass)** |

**Preparation time**: 20 mins

**Method**:

i. Weigh 30g of premix flour using weighing scale or scoop (30g scoop size) into a bowl and add little amount of water to make it into a slurry

ii. Transfer the slurry mixture into the boiling water

iii. Mix it thoroughly and cook for atleast 15 mins until the required consistency

iv. Further add a tsp of ghee , mix it and ready to serve

- **Recipe No.3: Idly**

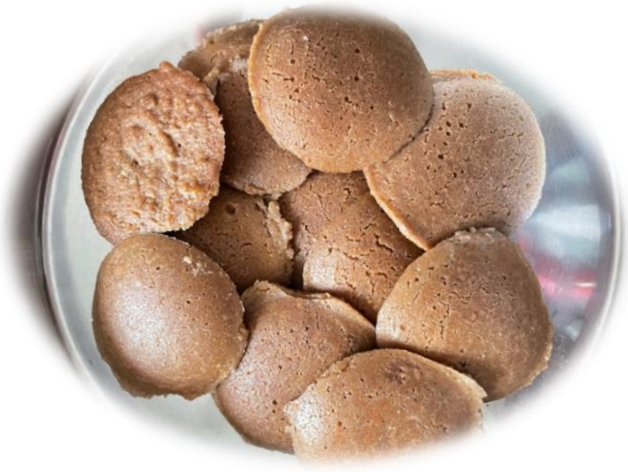

| **Ingredients** | **Quantity (g)** |
|---|---|
| **Premix powder** | **30** |
| **Curd** | **5 (1 tsp)** |
| **Water** | **As required** |

**Preparation time:** 20mins

**Method**:

i. Weigh 30g of premix flour using weighing scale or scoop (30g scoop size) into a bowl.
ii. Add curd (1 tsp) and required amount of water for batter consistency and keep it aside for at-least 1 hour.
iii. Keep the idly cooker ready with water and heat it.
iv. To the idly plates, apply little oil/ghee and pour in the batter.
v. Then arrange the idly plates in the cooker and cook it for 15 mins until it is perfectly steamed and serve it accordingly.

- **Recipe No.4: Dosa**

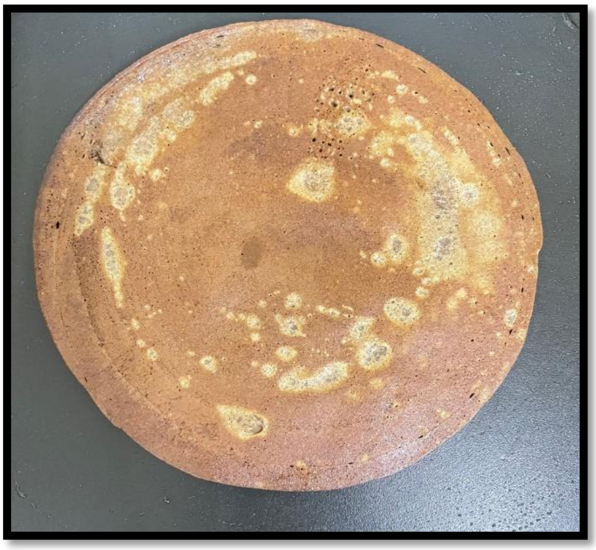

| **Ingredients** | **Quantity (g)** |
|---|---|
| Premix powder | 30 |
| Egg | 1 whole |
| Water | As required |

**Preparation time**: 15 mins

**Method**:

i. Weigh 30g of premix flour using weighing scale or scoop (30g scoop size) into a bowl.
ii. In a bowl, whisk one whole egg and transfer the premix into it.
iii. Then adjust the consistency by adding little amount of water and mix it properly.
iv. Keep it aside for at-least 1 hour.
v. Spread a ladleful of batter on to the preheated pan.
vi. Sprinkle little amount of oil/ghee and allow it cook on both sides.
vii. Once turned brown on both the sides and cooked properly, it is ready to serve.

### Feeding time table

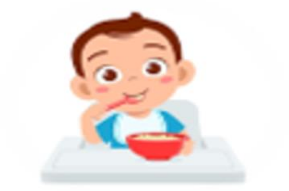

| Day | Mid-morning snack | Evening snack |
|---|---|---|
| Monday | Idly | Laddu |
| Tuesday | Porridge | Laddu |
| Wednesday | Dosa/ Idly | Laddu |
| Thursday | Porridge | Laddu |
| Friday | Dosa/ Idly | Laddu |

FINGER MILLET PROJECT

### Recipe Manual

Anganwadi Center code: ___________________________

Anganwadi Center Name: __________________________

Teacher's Name: __________________________________

INDIAN COUNCIL OF MEDICAL RESEARCH
NATIONAL INSTITUTE OF NUTRITION

**FOOD DIARY**

**Student Name:** **Centre Name:**

**UID** **Morning**

| **Date** | **Serving size (g)** | **Attendance (Yes/No)** | **Consumed (Yes/No)** | **Recipe Name** | **Given Quantity (g)** | **Left over (g)** | **Remarks** |
|---|---|---|---|---|---|---|---|

### FOOD DIARY

**Student Name:** **Centre Name:**

**UID:** **Evening**

| **Date** | **Serving size (g)** | **Attendance (Yes/No)** | **Consumed (Yes/No)** | **Recipe Name** | **Given Quantity (g)** | **Left over (g)** | **Remarks** |
|---|---|---|---|---|---|---|---|

**Adverse Events**

**Student Name:** **Centre Name:**

**UID:**

| **Date.** | **Physical Activity Level** | **Loss or reduced Appetite** | **Cough/Cold** | **Fever** | **Loose Motions** | | | **Constipation** | **Vomitings** |
|---|---|---|---|---|---|---|---|---|---|
| | **1. Normal 2. Sleepy 3. Difficult to awaken** | | | | **Frequency (Number)** | **with Blood** | **with Mucus** | | |

| Date | Bloated Stomach | Abdominal pain | Skin rashes | Other diseases, specify | Treatment | Any food allergy after consuming product | Food Allergy Response | | | Remarks |
|---|---|---|---|---|---|---|---|---|---|---|
| | | | | | Any Antibiotic or medication (Yes/No) | Yes/No If Yes specify | Immediate | <6 hours in a day | >6 hours in a aday | |
